# Loss and Reorganisation of Superficial White Matter in Alzheimer’s disease: A Diffusion MRI study

**DOI:** 10.1101/2021.05.07.21256728

**Authors:** Thomas Veale, Ian B. Malone, Teresa Poole, Thomas D. Parker, Catherine F. Slattery, Ross W. Paterson, Alexander J.M. Foulkes, David L. Thomas, Jonathan M. Schott, Hui Zhang, Nick C. Fox, David M. Cash

**Author notes:** Correspondence to: David Cash, Dementia Research Centre, UCL Queen Square Institute of Neurology, London, WC1N 3BG, UK.

## Abstract

Pathological involvement of cerebral white matter in Alzheimer’s disease has been shown using diffusion tensor imaging. Superficial white matter (SWM) changes have been relatively understudied despite their importance in cortico-cortical connections. Measuring SWM degeneration using diffusion tensor imaging is challenging due to its complex structure and proximity to the cortex. To overcome this we investigated diffusion MRI changes in young-onset Alzheimer’s disease using standard diffusion tensor imaging and Neurite Orientation Dispersion and Density Imaging to distinguish between disease-related changes that are due to degeneration (e.g. loss of myelinated fibres) and those due to reorganisation (e.g. increased fibre dispersion). Twenty-nine young-onset Alzheimer’s disease patients and 22 healthy controls had both single-shell and multi-shell diffusion MRI. We calculated fractional anisotropy, mean diffusivity, neurite density index, orientation dispersion index and tissue fraction (1-free water fraction). Diffusion metrics were sampled in 15 *a priori* regions of interest at four points along the cortical profile: cortical grey matter, the grey/white boundary, SWM (1mm below grey/white boundary) and SWM/deeper white matter (2mm below grey/white boundary). To estimate cross-sectional group differences, we used average marginal effects from linear mixed effect models of participants’ diffusion metrics along the cortical profile. The SWM of young-onset Alzheimer’s disease individuals had lower neurite density index compared to controls in five regions (superior and inferior parietal, precuneus, entorhinal and parahippocampus) (all *P*<0.05), and higher orientation dispersion index in three regions (fusiform, entorhinal and parahippocampus) (all *P*<0.05). Young-onset Alzheimer’s disease individuals had lower fractional anisotropy in the SWM of two regions (entorhinal and parahippocampus) (both *P*<0.05) and higher fractional anisotropy within the postcentral region (*P*<0.05). Mean diffusivity in SWM was higher in the young-onset Alzheimer’s disease group in the parahippocampal region (*P*<0.05) and lower in three regions (postcentral, precentral and superior temporal) (all *P*<0.05). In the overlying grey matter, disease-related changes were largely consistent with SWM findings when using neurite density index and fractional anisotropy, but appeared at odds with orientation dispersion and mean diffusivity SWM changes. Tissue fraction was significantly lower across all grey matter regions in young-onset Alzheimer’s disease individuals (all *P*<0.001) but group differences reduced in magnitude and coverage when moving towards the SWM. These results show that microstructural changes occur within SWM and along the cortical profile in individuals with young-onset Alzheimer’s disease. Lower neurite density and higher orientation dispersion suggests underlying SWM fibres undergo neurodegeneration and reorganisation, two effects previously indiscernible using standard diffusion tensor metrics in SWM.

## Introduction

Alzheimer’s disease is characterised by progressive neurodegenerative changes that involve both grey matter (GM) and white matter (WM). Macrostructural GM loss as visualised in hippocampal atrophy using MRI is characteristic of Alzheimer’s disease.^1,2^ However, microstructural change may appear before macrostructural neurodegeneration and occur in early stages of the disease.^3–5^ Disconnection of neural circuits by means of WM disruption is likely a key driver for cognitive deficits and may be an important pathophysiological process of Alzheimer’s disease.^6,7^

WM is heterogeneous and can be broadly divided into superficial white matter (SWM) and deep white matter (DWM).^8,9^ SWM mainly consists of short, thin, U-fibres linking nearby gyri that lie up to 2mm below the cortex but may represent 57-67% of all WM fibres.^10–12^ SWM fibre organisation is also highly complex^13,14^ and may form sub-networks of its own.^15^ Unique anatomical, developmental and cellular SWM properties may lead to particular vulnerabilities to Alzheimer’s disease-related pathology. For example, short, thin fibres prevalent in SWM are the last WM outside the cortex to myelinate,^16,17^ which means oligodendrocytes in these regions may be more vulnerable to metabolic insults.^18,19^ SWM contains the highest density of interstitial cells in WM that harbour neurofibrillary tangles.^20,21^ Amyloid-β is also deposited in the SWM of humans and qualitatively associated with focal SWM demylination in mice.^22,23^

Although SWM may be vulnerable to Alzheimer’s disease pathology and represents the majority of cerebral WM, relatively few studies have investigated *in vivo* SWM changes in Alzheimer’s disease. Magnetisation transfer ratio studies sampled at 3mm below the GM/WM boundary found those with Alzheimer’s disease show demyelination in SWM.^24,25^ SWM diffusion MRI (dMRI) studies in Alzheimer’s disease and mild cognitive impairment (MCI) have shown increased axial, radial and mean diffusivity (MD) in temporal, parietal and occipital regions that are associated with changes in the Mini Mental State Examination (MMSE) and with increased reaction time on a prospective memory task.^26–29^

SWM’s proximity to cortical GM and its complicated fibre organisation present two major methodological challenges that hinder the ability of neuroimaging to localise disease-related changes specific to SWM. Firstly, relatively large dMRI voxel sizes and varying sampling distances below the GM/WM boundary make it unclear whether changes attributed to SWM arise from the cortical GM, SWM or DWM. Secondly, dMRI signal is often modelled using a single tensor that may not capture the organisationally complex microstructure of SWM. Consequently Alzheimer’s disease-related SWM changes in commonly used DTI metrics, such as fractional anisotropy (FA) and MD, may result from a mixture of WM degeneration and reorganisation.^30^ Therefore, current studies showing Alzheimer’s disease-related changes in SWM may be due to an unknown combination of disease-related effects and confounding methodological issues.

In order to address some of the challenges with quantifying disease-related changes in SWM, we used Neurite Orientation Dispersion and Density Imaging (NODDI) to disentangle neurodegenerative and organisational alterations by independently quantifying the neurite density index (NDI) and orientation dispersion index (ODI) of underlying SWM fibres.^31^ NODDI metrics relate to histological measures of myelin and tau pathology.^32–34^ Alzheimer’s disease-related changes in the GM and DWM have also been observed in those with young-onset Alzheimer’s disease.^35,36^ The NODDI model also allows separation of free water fraction (FWF) and brain tissue, enabling the removal of the confounding contribution of nearby CSF. We sampled NODDI and DTI measures at various depths along the cortical profile (beginning in cortical GM and descending into the SWM) in 15 *a priori* cortical regions of interest (ROI) to better understand effects of Alzheimer’s disease on SWM and its neighbouring tissue. We hypothesised that we would replicate significant Alzheimer’s disease-related changes in SWM using standard DTI metrics, and that significant Alzheimer’s disease-related SWM changes would remain when modelling the density and dispersion of neurites using NODDI.

## Materials and Methods

### Participants and Exclusions

Sixty-nine participants were recruited, 45 with young-onset Alzheimer’s disease and 24 healthy controls. The young-onset Alzheimer’s disease patients were recruited from a specialist Cognitive Disorders Clinic between 2013 and 2015 with probable Alzheimer’s disease^37^ and symptom onset < 65 years old. Twenty-eight patients were classified as having typical (amnestic onset) Alzheimer’s disease and 17 with atypical Alzheimer’s disease, 14 of whom were diagnosed with posterior cortical atrophy. Twenty-four cognitively healthy controls with similar mean age and sex to the Alzheimer’s disease cohort were recruited. This cohort has been described in Slattery *et al*^36^ and Parker *et al*.^35^ Each participant underwent MRI and cognitive tests including Mini Mental State Examination (MMSE); see Slattery *et al*^36^ for details of cognitive assessments. Ethical approval was obtained from the National Hospital for Neurology and Neurosurgery Research Ethics Committee and written informed consent was obtained from all participants.

Participants were only included in analysis if they had suitable quality T1w, single-shell DTI and multi-shell NODDI data (see *MRI Acquisition*, below). From the original 69 participants, one participant was excluded due to excessive T1w motion, three due to image processing failures of T1w images, seven due to motion and processing failures in DTI sequences and seven due to motion and processing failures in NODDI sequences. After exclusions, 51 participants remained for analysis (Table 1).

**Table 1:** Demographics of Analysed Participants.

|  | Control (n = 22) | Young-Onset Alzheimer's disease (n = 29) |
| --- | --- | --- |
| Females/Males (n) | 12/10 | 16/13 |
| Age (SD) | 60.5 (5.7) | 61.7 (4.6) |
| Age at Onset (SD) | N/A | 56.6 (4.2) |
| MMSE median (IQR) | 30 (1)<sup>a*</sup> | 21 (7)<sup>a*</sup> |
| Cortical Thickness (SD) | 2.59 (0.08)<sup>b*</sup> | 2.49 (0.12)<sup>b*</sup> |

<sup>a</sup>Mann-Whitney test.
<sup>b</sup>Welsch's t-test.
\* indicates $p < 0.001$ .
All data are mean (SD) unless stated otherwise. Cortical thickness (mm) is averaged across *a priori* ROIs for each participant in each group. Abbreviations: n = number; MMSE = Mini-Mental State Examination; SD = standard deviation; IQR = interquartile range.

### MRI Acquisition

MR images were acquired on a Siemens Magnetom Trio (Siemens, Erlangen, Germany) 3 Tesla MRI with a 32-channel phased array receiver head coil. Structural MRI sequences included sagittal 3D MPRAGE T1w volumetric MRI (MPRAGE with TE/TI/TR = 2.9/900/2200 ms, matrix size 256 ⨯ 256 ⨯ 208, voxel size 1.1mm isotropic) and 3D T2w volumetric MRI (SPACE with TE/TR = 401/3200ms, matrix size 256 x 256 x 176, voxel size 1.1mm isotropic). Diffusion MRI sequences included a dedicated single-shell acquisition for DTI (spin echo EPI; 64 diffusion-weighted directions; b = 1000 s/mm^2^; 9 b=0 s/mm^2^ images; 96 ⨯ 96 ⨯ 55 slices; voxel size 2.5mm isotropic; TR/TE = 6900/91 ms) and a multi-shell sequence optimised for NODDI (spin echo EPI; 64, 32, and 8 diffusion-weighted directions at b=2000, b=700, and b=300 s/mm^2^; 13 interleaved b=0 s/mm^2^; 96 ⨯ 96 ⨯ 55 slices; voxel size 2.5mm isotropic; TR/TE = 7000/92 ms). B_0_ field maps for single-shell and NODDI sequences were acquired separately to correct for susceptibility-related distortion in the diffusion images (TE 1,2/TR = 4.92,7.38/688ms, 64 ⨯ 64 ⨯ 55, voxel size 3mm isotropic). DTI and NODDI sequences were acquired across 2 scanning sessions (mean scan interval = 8.7 days; SD = 11.2). Participant ages were calculated at their NODDI scan.

### Image Processing

Structural images were processed to obtain a high-resolution reconstruction of the cortical surface. The T1w image was first skull-stripped using a brain mask from Geodesic Information Flow (GIF).^38^ Cortical surface reconstruction was then performed on the skull-stripped image using FreeSurfer 6.0 (http://surfer.nmr.mgh.harvard.edu/) to segment GM and WM, tessellate the GM/WM boundary, and to perform automated topology correction and surface deformation for optimisation of the GM/WM boundary and GM/CSF boundary.^39,40^ A volumetric T2w image was available in all but one participant and included in the FreeSurfer pipeline in order to improve detection of the pial surface. After reconstruction, cortical thickness values were extracted for ROIs based on the the Desikan-Killiany atlas.

Diffusion-weighted image processing involved skull-stripping with a total intracranial volume mask using SPM12,^41^ motion and eddy-current correction using FSL’s eddy tool^42^ and susceptibility correction using a combined approach of phase unwrapping and registration.^43^ NiftyFit^44^ was used to fit a DTI model to the single-shell data with a weighted-least squares approach, and MD and FA maps were extracted from resulting tensor images. NODDI measures were fitted using Accelerated Microstructural Imaging via Convex Optimisation (AMICO),^45^ which is a linearized formulation of the NODDI model that tends to improve the speed and stability of the fit in regions near the cortex.^45,46^ From the model, three measures were produced at each voxel: NDI, ODI and FWF. NDI and ODI quantify the density and orientation dispersion of neurites in the tissue fraction (TF) of the voxel (where TF = 1-FWF). FA, MD, NDI, ODI and TF maps were used in the analysis of the cortical profile.

DTI and NODDI maps were registered to the T1w image with NiftyReg^47^ using the first b=0 of each sequence. The resulting transformations were used to resample the DTI and NODDI metrics into T1w space using cubic interpolation. All resampled dMRI images were visually reviewed to ensure accurate alignment with the FreeSurfer GM/WM boundary.

### Cortical Profile Extraction

dMRI metrics were sampled along the cortical profile to allow greater localisation of pathological changes in SWM compared to neighbouring tissues. For each vertex on the surface representing the GM/WM boundary, DTI and NODDI metrics were sampled at four distances from the vertex along the surface normal: 1mm outwards (likely GM), 0mm (GM/WM boundary), 1mm inwards (likely SWM), and 2mm inwards (likely a mixture of SWM and DWM).

### Regions of Interest (ROIs) Measures

For each of the four points sampled along the cortical profile, summaries of each dMRI metric were obtained on a region of interest (ROI) level based on anatomical labels from the FreeSurfer Desikan-Killiany atlas brain parcellation. A total of 15 *a priori* ROIs were included in the analysis. Twelve were chosen from temporal, occipital and parietal regions that are known to be affected in both clinical phenotypes within the young-onset Alzheimer’s disease group (typical Alzheimer’s disease and posterior cortical atrophy)^48,49^: entorhinal, superior temporal, fusiform, lateral occipital, middle temporal, posterior cingulate, inferior parietal, parahippocampal, cuneus, inferior temporal, precuneus and superior parietal cortices. Three ROIs in somatosensory cortices, typically spared until later stages of typical Alzheimer’s disease and posterior cortical atrophy,^48,49^ were chosen to serve as control ROIs: precentral, postcentral and paracentral cortices. ROI measures for DTI and NODDI metrics were averaged across hemispheres. See Fig. 1 for the full preprocessing pipeline.

**Figure 1:**
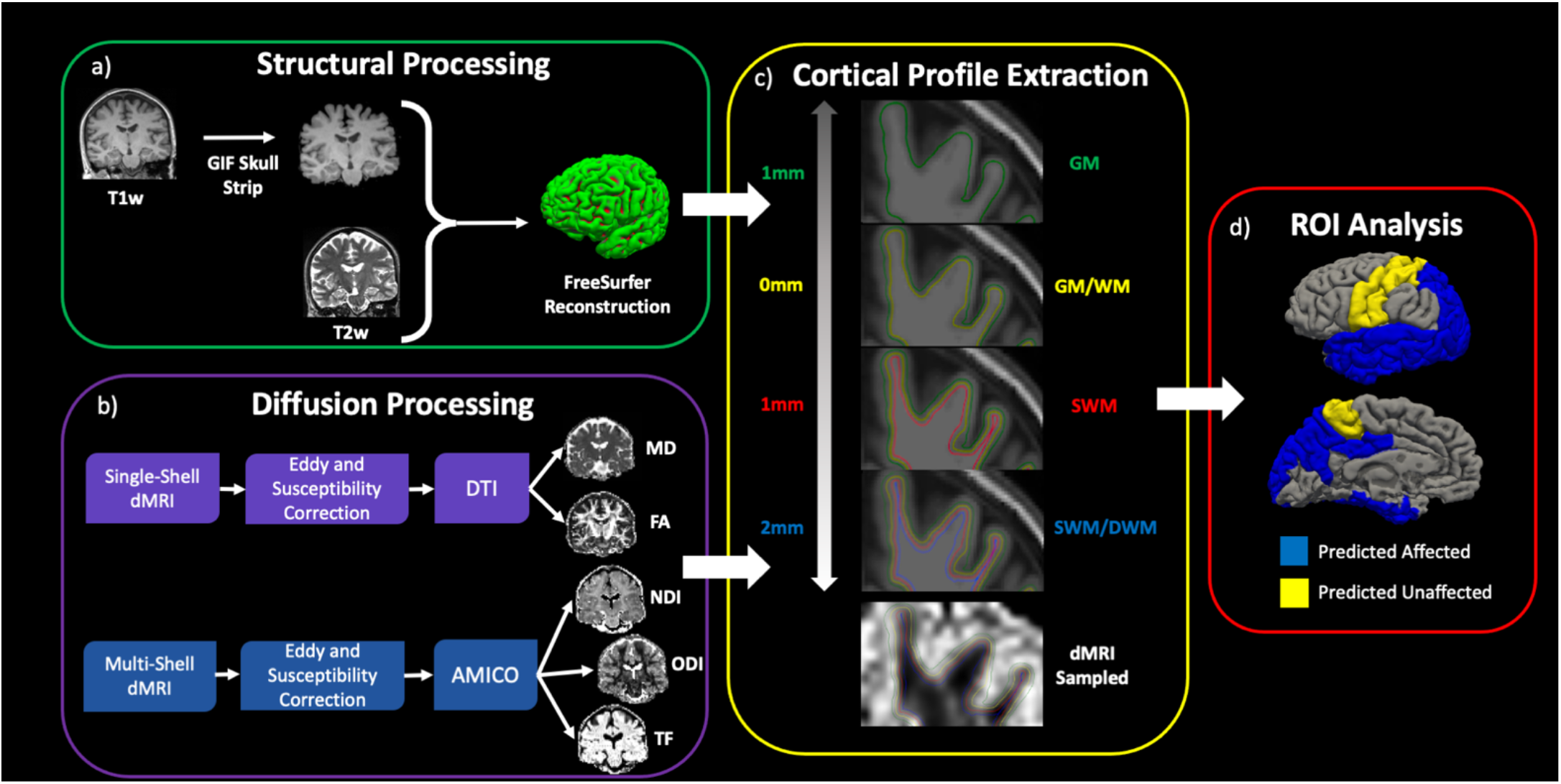
Schematic of preprocessing pipeline to acquire ROI-level metrics at varying levels from the GM/WM boundary. **a)** Structural MRI processing for skull stripping and surface reconstruction to define the GM/WM boundary. **b)** dMRI processing to create DTI maps (MD and FA) and NODDI maps (NDI, ODI and TF). **c)** dMRI metrics are sampled at various distances from the GM/WM boundary: GM (1mm outwards in green), GM/WM (on the boundary in yellow), SWM (1mm inwards in red) and SWM/DWM (2mm inwards in blue). **d)** ROI measures of dMRI metrics, sampled at each distance, are extracted using the Desikan-Killiany atlas from FreeSurfer. ROIs were hypothesised to either be affected (blue) or not affected (yellow) in the young-onset Alzheimer’s disease group.

While NDI and ODI measures are estimated within the TF of each voxel, a conventional ROI average would equally weight NDI and ODI across the region despite varying TF likely to occur near the cortex. Therefore, we used “tissue-weighted” averages of NDI and ODI for all analyses, where TF at corresponding vertices served as the weights.

### Statistical Analysis

All statistical analyses were performed using R 3.6.3^50^ and ROI visualisations were plotted using the *ggseg* package.^51^ In addition to plotting individual ROI measures from our main analysis, we averaged the observed dMRI metrics across all 15 *a priori* ROIs to obtain a composite overview across the four distance points of the cortical profile (Fig. 2).

**Figure 2:**
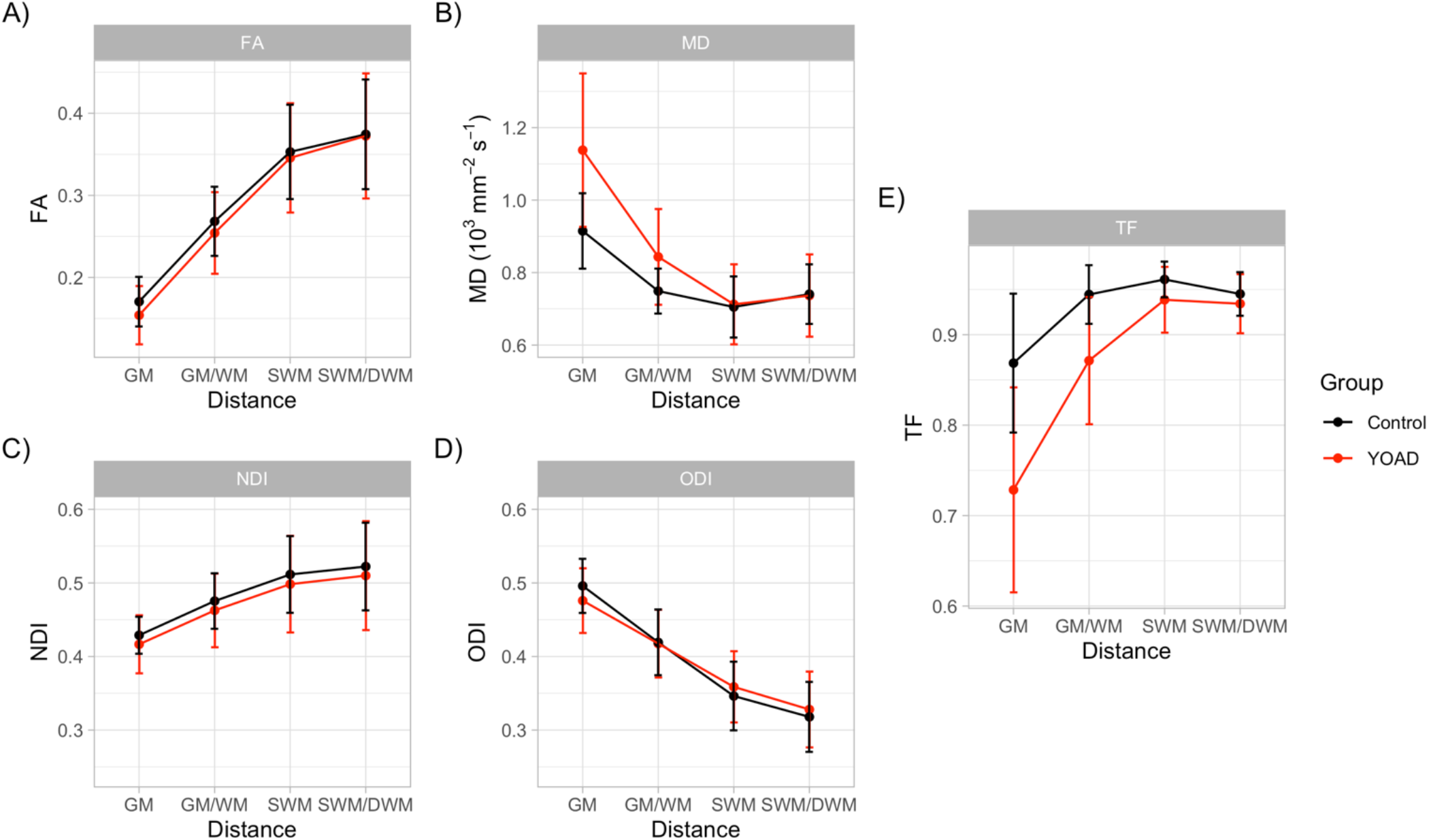
Average observed dMRI metrics (over all *a priori ROIs)* across the GM/WM boundary for both controls (black) and young-onset Alzheimer’s disease (red) groups. **A)** Averaged FA profile shows an increase for both groups when moving into SWM/DWM. **B)** Average MD profile shows an overall decrease when moving into the SWM/DWM. **C)** Average NDI profile shows an overall increase when moving into SWM/DWM. **D)** Average ODI profile shows an overall decrease when moving into SWM/DWM. **E)** For reference, the average TF profile shows an overall increase when moving into SWM/DWM. Error bars indicate +/- standard deviation across all 15 ROIs, for each group, at each distance from GM/WM boundary. YOAD = Young-Onset Alzheimer’s disease.

Disease-related changes in dMRI metrics along the cortical profile were modelled using linear mixed effect models. A total of 75 models were generated (15 *a priori* ROIs ×five dMRI metrics). In each model, the outcome variable was the ROI average of the dMRI metric. The model included fixed effects for distance, distance-squared, group (control or young-onset Alzheimer’s disease), average cortical thickness of the ROI, plus interactions between distance and group, and distance-squared and group terms (see Supplementary Material for equation). Quadratic term for distance was used to capture non-linear trends observed when plotting observed data for each ROI (Fig. 2). Cortical thickness was included in the model as a proxy for atrophy and we assumed this would be most associated with diffusion measures in the GM. We therefore set the GM point as the intercept for the model (distance=0), with distance increasing in the direction towards the SWM/DWM (distance=4). Age and sex were not included in the model as they were well balanced in our cohort and including these as covariates produced no meaningful change in results across our models. Group-specific random effects were included for the intercept and slope, along with a covariance term between the intercept and slope. In a small number of models (6/75) where there was difficulty fitting this model, we removed the random slope for the control group (see Supplementary Material). The average marginal effect (AME) of group, at each of the four distance points, was estimated using predicted values from the linear mixed effect models. AMEs represent the estimated difference in the dMRI metric between the young-onset Alzheimer’s disease group compared with controls, using the observed values of other covariates (here, the cortical thickness). Multiple comparisons were corrected for using the False Discovery Rate (FDR), where corrected *P*-values were considered statistically significant below a threshold of 0.05 (*pFDR* < 0.05).^52^ Residuals for each model were inspected to check model assumptions were adequately met. We also estimated AMEs across the GM/WM boundary for NODDI measures when using a conventional mean rather than tissue-weighted mean (Supplementary Table 4; Supplementary Fig. 5-6). Example R code for fitting linear mixed effect models and extracting AMEs is included in the supplementary material.

### Data Availability

An anonymised spreadsheet with data used to fit linear mixed effect models along the cortical profile can be made available upon reasonable request from researchers.

## Results

Fig. 2 shows the observed DTI and NODDI metrics averaged across 15 *a priori* ROIs along the cortical profile. When using DTI metrics, there was an overall increase in FA when moving from the GM into the SWM/DWM (Fig. 2A) and an overall decrease in MD (Fig. 2B) when moving from the GM into the SWM and DWM. When using NODDI metrics, there was an overall increase in NDI when moving from the GM into the SWM/DWM (Fig. 2C) and an overall decrease in ODI when moving from the GM into the SWM/DWM (Fig. 2D). There was an overall increase in TF when moving from GM into SWM/DWM (Fig. 2E).

Group differences, at each of the four distance points across the GM/WM boundary, were then investigated for each ROI (Fig. 3A) and each of the five metrics, using AMEs and their 95% confidence intervals (all reported results significant at *pFDR* < 0.05). See Supplementary Fig. 1-3 and Supplementary Table 1-3 for all AMEs.

Statistically significant findings across the cortical profile using DTI metrics are shown in Fig. 3B-C. In the SWM (1-2mm below the GM/WM boundary), those with young-onset Alzheimer’s disease had significantly lower FA compared to controls in the parahippocampal (SWM: *AME* -0.026 [95% *CI*: -0.039, -0.013]; SWM/DWM: -0.021 [-0.037, -0.005]) and entorhinal SWM (SWM: -0.031 [-0.045, -0.016]), but higher FA within the postcentral SWM (SWM/DWM: 0.028 [0.010, 0.047]). At the GM and on the GM/WM boundary, young-onset Alzheimer’s disease individuals had significantly lower FA in six ROIs: parahippocampal, entorhinal, inferior and superior parietal, lateral occipital cortices and the cuneus (Fig. 3B).

**Figure 3:**
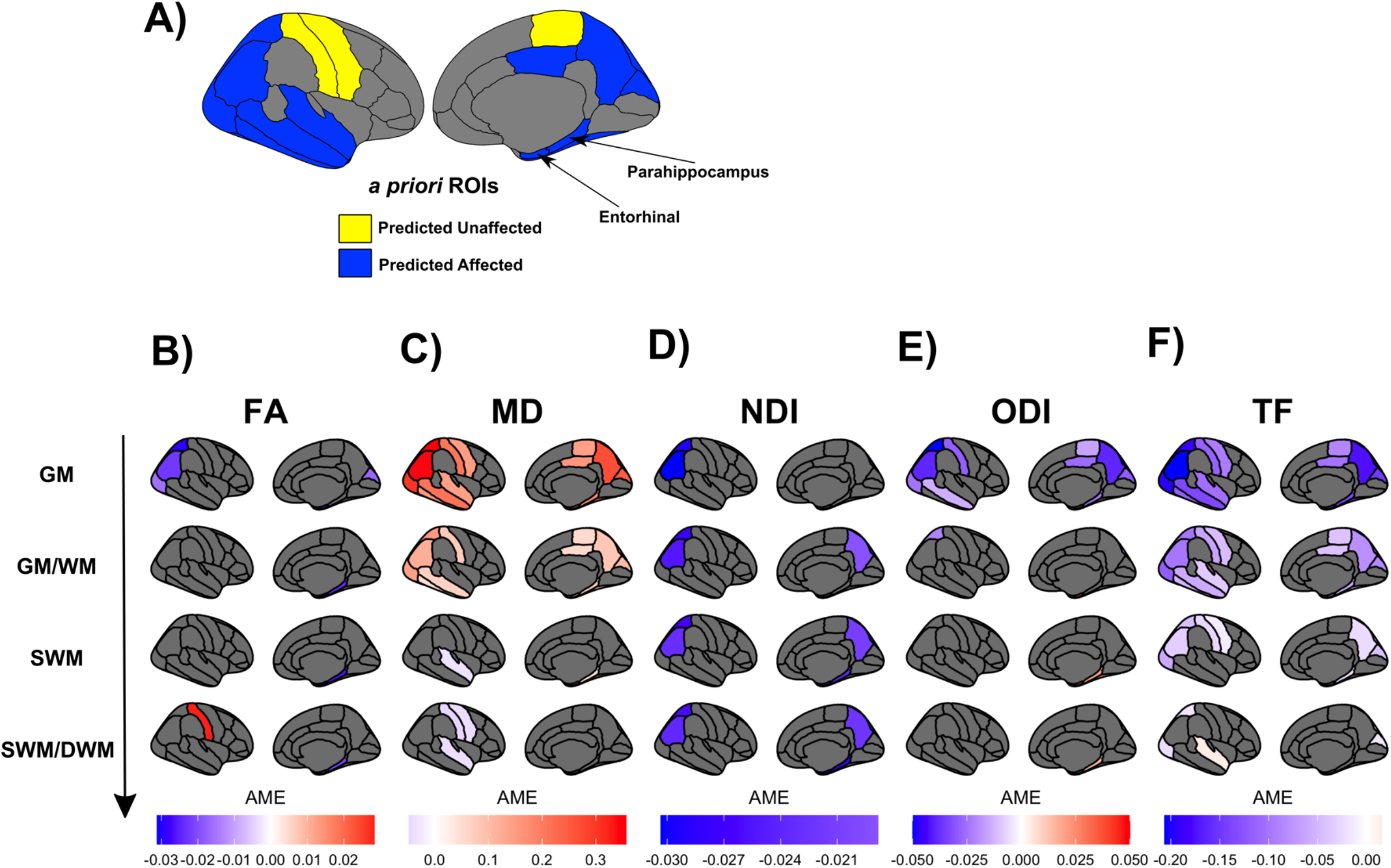
Regional group differences along the cortical profile. **A)** *A priori* ROIs were chosen due to prior knowledge of affected areas in young-onset Alzheimer’s disease (blue) or as control ROIs (yellow). Diffusion metrics were obtained along the cortical profile. **B-F)** DTI, NODDI and TF Average Marginal Effects (AMEs) (*pFDR* < 0.05) representing the difference in dMRI metric between controls and young-onset Alzheimer’s disease, in each ROI, at each distance across the GM/WM boundary from the linear mixed effect models (GM (1mm outward from GM/WM boundary), GM/WM boundary, SWM (1mm inwards from GM/WM boundary), SWM/DWM (2mm inwards from GM/WM boundary)). Negative AMEs (purple/blue colourmap) indicates a lower dMRI metric in the young-onset Alzheimer’s disease group, while positive AMEs (red/cream colourmap) indicate a higher metric in the young-onset Alzheimer’s disease group. *A priori* ROIs not coloured in plots have *pFDR* > 0.05. Supplementary Fig. 4 shows AMEs at uncorrected *P* < 0.05.

The young-onset Alzheimer’s disease group showed significantly higher MD compared to controls in the parahippocampal SWM (SWM: 0.038 [0.009, 0.067]), but significantly lower MD in the SWM of pre- and postcentral gyri, and the superior temporal region. This contrasts with young-onset Alzheimer’s disease individuals having significantly higher MD across all ROIs in the GM and in all but two ROIs in on the GM/WM boundary (Fig. 3C).

Statistically significant findings across the cortical profile using NODDI metrics are shown in Fig. 3D-E. In the SWM (1-2mm below the GM/WM boundary), individuals with young-onset Alzheimer’s disease had significantly lower NDI in the inferior and superior parietal, precuneus, parahippocampal and entorhinal regions (Fig. 3D). In parietal regions, lower NDI in those with young-onset Alzheimer’s disease was consistent with lower NDI the overlying GM and on the GM/WM boundary, but with no significant group differences were observed within entorhinal, parahippocampal and precuneus GM.

Young-onset Alzheimer’s disease individuals had significantly higher ODI compared to controls in the SWM of the parahippocampal (SWM: 0.020 [0.010, 0.030]; SWM/DWM: 0.016 [0.003, 0.030]), entorhinal (SWM: 0.033 [0.019, 0.048]) and fusiform ROIs (SWM: 0.017 [0.004, 0.030]). This largely contrasted with findings in the GM, where those with young-onset Alzheimer’s disease had significantly lower ODI in widespread regions of GM such as parahippocampal, superior and inferior parietal, middle temporal and posterior cingulate ROIs. On the GM/WM boundary, those with young-onset Alzheimer’s disease also had lower ODI compared to controls in the superior parietal lobe (GM/WM: -0.022 [-0.035, - 0.008]) but significantly higher ODI in the entorhinal ROI (GM/WM: 0.029 [0.016, 0.042]) (Fig. 3E).

We report statistically significant group differences for TF along the cortical profile to provide a reference for DTI and NODDI metric results with respect to macroscopic diffusion changes (Fig. 3F). TF in GM and on the GM/WM boundary was significantly lower in young-onset Alzheimer’s disease compared to controls across all *a priori* ROIs. TF in SWM remained significantly lower in young-onset Alzheimer’s disease compared to controls, but in fewer ROIs and at a reduced magnitude. TF in SWM/DWM was significantly lower in young-onset Alzheimer’s disease compared to controls in only three ROIs but higher within the superior temporal ROI (SWM/DWM: 0.016 [0.005, 0.026]) (Fig. 3F).

## Discussion

In this study we show that 1) NODDI detects disease-related microstructural changes in the SWM of those with young-onset Alzheimer’s disease; 2) these Alzheimer’s disease-related SWM changes do not wholly overlap with DTI metrics; and 3) these microstructural changes vary in their relationship to overlying GM. To our knowledge, this work is both the first to investigate Alzheimer’s disease-related SWM changes using NODDI and to contextualise these findings within the cortical profile of individuals with Alzheimer’s disease.

Our main findings indicate that individuals with young-onset Alzheimer’s disease have reduced NDI in the SWM of parietal, parahippocampal, entorhinal and precuneus SWM but increased ODI in the parahippocampal, entorhinal and fusiform SWM. These effects appear to overlap in some regions (e.g. parahippocampal SWM). However other regions may only detect either a loss of fibres indicated by decreased NDI (e.g. precuneus SWM) (Fig. 3D), or fibre reorganisation indicated by increase ODI (e.g. fusiform SWM) (Fig. 3E). This suggests that myelinated fibres in the SWM undergo both neurodegeneration and reorganisation in young-onset Alzheimer’s disease but that these changes may not always co-localise.

NODDI SWM findings overlap with conventional DTI metrics in regions typically affected early in the disease such as the parahippocampus and entorhinal regions.^53,54^ For example, those with young-onset Alzheimer’s disease had lower NDI and FA, but higher MD and ODI, in parahippocampal SWM compared to controls (Fig. 3B-3E). A simultaneous reduction in NDI and increase in ODI here implies FA and MD changes are being driven by both degeneration and reorganisation of WM fibres. These concomitant decreases in FA and NDI with increases in ODI have also been observed in the WM of transgenic mice models of tauopathy that are linked to WM disorganisation.^32,55^ Moreover, interstitial cells prevalent in SWM are susceptible to cytoskeletal changes in the entorhinal region and may contribute to these disease-related SWM changes.^20^

In other regions, NODDI and DTI findings in SWM were not always congruent. Only NDI showed Alzheimer’s disease-related SWM changes in the parietal and precuneus regions, which likely reflects loss of underlying myelinated fibres but may not result in a detectable change of FA,^31,33^ potentially due to complex crossing fibres in the underlying SWM.^13–15^ A significant increase in ODI alone (e.g. fusiform SWM) indicates increased fibre dispersion in the young-onset Alzheimer’s disease group being the dominating microstructural change, with no statistically significant loss of underlying myelinated fibres. Myelinated fibre disorganisation has also been observed at post-mortem in the cortex of young-onset Alzheimer’s disease individuals^56^ but it is unknown if this extends into SWM.

Conversely, only DTI metrics indicated significant group changes in the superior temporal and central sulcus regions (Fig. 3B-3C). Higher FA and lower MD in young-onset Alzheimer’s disease individuals’ SWM is less clear, but one explanation could be compensatory mechanisms, particularly in regions typically affected by atrophy in later stages such as two of our control regions within the central sulcus. For example, previous FA increases have been observed in asymptomatic amyloid positive individuals^57^ and transgenic mouse models of Alzheimer’s disease prior to intraneuronal plaque accumulation.^58^ Alternatively, previous work has attributed increased anisotropy to a degeneration of crossing fibres and sparing of motor-related projection fibres.^59^ However, with no corresponding NDI or ODI changes in these regions that met our our FDR-corrected threshold for multiple comparisons, the specific microstructural changes driving DTI here are unclear.

Our results do not replicate previous DTI-based findings in the SWM of those with Alzheimer’s disease. Previous studies of Alzheimer’s disease-related changes in SWM found widespread increases in MD,^26,28,29^ where our results suggest higher MD in those with Alzheimer’s disease only within the parahippocampal SWM. We also report reduced MD in those with Alzheimer’s disease in the SWM of the central sulcus and superior temporal regions. This could be due to the inclusion of cortical thickness as a covariate in our models, reducing the influence of overlying GM atrophy on SWM metrics (see widespread increased MD in those with Alzheimer’s disease stopping on the GM/WM boundary in Fig. 3C). Taking the NODDI and DTI SWM findings together, NODDI metrics in SWM can detect distinct Alzheimer’s disease-related changes, but together with DTI depict a complex microstructural environment that is likely undergoing spatially heterogeneous pathophysiological affects of Alzheimer’s disease. Further investigation into Alzheimer’s disease-related SWM changes is needed to determine the source of these microstructural effects.

Diffusion metrics also varied when moving along the cortical profile for all participants (Fig. 2). FA increased and MD decreased when moving from GM into SWM, which suggests diffusion in the SWM and DWM is more restricted and anisotropic than in the GM. Kang *et al*.^60^ also showed increasing FA and decreasing MD in healthy controls and suggested MD values were significantly affected by partial voluming of CSF. Cortical atrophy in the young-onset Alzheimer’s disease group would increase the likelihood of CSF contamination within voxels classified as GM, making it difficult to determine whether changes are entirely due to intrinsic tissue properties from FA and MD alone. Conversely, NODDI metrics allow for the modelling of this partial volume effect with the free water compartment.^31^ While explicitly taking free water into account on the regional level using tissue-weighted averages, NDI increased and ODI decreased when moving from GM into the SWM which provides evidence that changes in FA and MD are partially due to histological features of increased myelination and parallel fibre organisation in WM.

When viewing group effects with NODDI along the cortical profile, young-onset Alzheimer’s disease individuals showed a sustained reduction in NDI in regions such as the precuneus compared to controls, which may indicate reduced density of myelinated neurites in both GM and SWM.^33^ Young-onset Alzheimer’s disease individuals showed decreased ODI within GM but increased ODI within certain SWM regions compared to controls. Although most prominent in the parahippocampus, there is a trend of higher ODI in young-onset Alzheimer’s disease individuals across more SWM regions when uncorrected for multiple comparisons (Supplementary Fig. 4). A potential flattening of ODI along the cortical profile suggests neurite organisation could become more similar across the GM/WM boundary in young-onset Alzheimer’s disease and may be associated with blurring of the GM/WM boundary observed in T1w sequences.^61–63^

dMRI measures in GM and SWM are likely affected by confounding effects of nearby CSF which is further implicated by GM atrophy in our young-onset Alzheimer’s disease group. We decided to account for CSF partial volume effects using 1) cortical thickness measures within the models for all dMRI metrics and 2) calculating NODDI metrics as tissue weighted averages where TF acted as the weights (where TF = 1-FWF). We also analysed TF changes in order to determine whether macrostructural diffusion changes, likely attributed to the influence of nearby free water in CSF, are occuring simultaneously in these regions across the cortical profile (Fig. 3F). The young-onset Alzheimer’s disease group had lower TF across all GM regions with smaller but significant group differences remaining in the SWM of fewer regions. This suggests that disease-related changes in diffusion tensor metrics in the GM and on the GM/WM boundary are likely to be strongly influenced by free water and partial volume effects. Indeed, striking similarities between MD and TF measures can be observed in the cortical profiles and group differences (Fig. 2B and 2E; Fig. 3C and 3F). Exploratory scatterplots of MD vs TF indicates a prominent negative relationship in the GM and on the GM/WM boundary across all *a priori* ROIs, but diminishes when entering SWM (Supplementary Fig. 7). Therefore MD findings in the GM and GM/WM boundary may be driven by FWF in the nearby CSF. This suggests previous Alzheimer’s disease-related SWM changes sampled on the GM/WM boundary may be influenced by partial volume effects and supports findings of CSF confounding DTI measures in Alzheimer’s disease that can artificially inflate MD.^64^ Due to SWM’s proximity to the overlying cortical GM we recommend the use of dMRI models that take free water into account, such as NODDI or free-water elimination DTI,^65,66^ to allow for more specific insights into pathological microstructural changes.

Nevertheless, when taking taking TF into account using tissue-weighted averages, NDI and ODI still showed disease-related changes in the GM. This complements previous NODDI work in GM of Alzheimer’s disease participants^35,67^ by showing Alzheimer’s disease-related reductions in NDI and ODI remain, even when explicitly accounting for the varying degree of FWF within regions. We also provide NODDI results with conventional regional averages as a reference point to previous GM work (Supplementary Fig. 5-6; Supplementary Table 4). Future studies could further overcome CSF and atrophy confounds by studying microstructural GM and SWM changes in individuals at-risk of developing Alzheimer’s disease before macrostructural neurodegeneration occurs.

Varying regional differences in NODDI and DTI metrics may also be due to the spatial distribution of underlying pathology of clinical phenotypes in our young-onset Alzheimer’s disease cohort (typical Alzheimer’s disease and posterior cortical atrophy) in addition to varying SWM properties across regions. For example, atrophy patterns do not completely overlap between typical Alzheimer’s disease (temporo-parietal atrophy) and posterior cortical atrophy (occipito-parietal atrophy),^49,68^ while the ability to detect reproducible *in vivo* SWM fibres using dMRI varies across regions and techniques.^9^ This spatial heterogeneity in both pathology and dMRI’s sensitivity to underlying SWM may obscure our ability to detect underlying microstructural changes. More work is needed to distinguish other contributing factors to *in vivo* SWM measures and how these would affect potential biomarkers for Alzheimer’s disease.

A limitation of this work is the potential influence of partial volume effects due to the spatial resolution of our dataset (2.5mm isotropic). We opted for a ‘WM mesh’ approach^9^ to determine Alzheimer’s disease-related changes within the SWM region as opposed to extracting specific U-fibre tracts. In addition, we sampled at four points across the GM/WM boundary and modelled the relationship between these points to mitigate misinterpetations of SWM changes. Despite these efforts some influence from partial volume effects will occur. Although U-fibres have been extracted at 1.25mm and 2mm isotropic dMRI resolutions^9,69^, submillimetre diffusion MRI greatly improves detection of U-fibres^70^. Indeed, recent advances in submillimetre dMRI enable highly reproducible *in vivo* U-fibre tracts^71^ and the ability to delineate SWM from nearby GM and DWM using iron levels in high-resolution quantitative MRI.^12^ This highlights the importance of high-resolution techniques for studying SWM *in vivo* and evaluating its trade-off with scan duration to determine the clinical applications of SWM-based biomarkers.

Another potential limitation is the role of cortical topography. Curvature of the cortex is known to influence dMRI measures, thus averaging dMRI metrics in an ROI across gyri and sulci could blur and mask true changes. Curvature also varies across brain regions throughout the lifespan and sulcal widening is a prominent feature of macrostructural Alzheimer’s disease changes, which may further contribute to heterogeneous SWM dMRI metrics.^72–74^ This could be investigated by segmenting ROIs into curvature-based subregions to determine if SWM measures vary in gyri versus sulci.^75^ Finally, SWM extraction using FreeSurfer is suboptimal as projecting along the surface normal from the GM/WM boundary can produce unrealistic SWM sampling. A recently developed gyral coordinate system that interpolates underlying fibre orientations from gyral morphology has shown to align with primary DTI eigenvectors, and may provide a novel way to investigate *in vivo* SWM changes in Alzheimer’s disease.^76^

In conclusion, we show that Alzheimer’s disease-related microstructural changes occur within SWM and along the cortical profile. We independently quantified the density and dispersion of underlying SWM neurites using NODDI, two metrics that cannot be disentangled using standard tensor metrics used in previous dMRI studies of SWM in Alzheimer’s disease. For the first time we show lower NDI and higher ODI in the SWM of those with young-onset Alzheimer’s disease, likely due to a loss of myelin and reorganisation respectively. Complex fibres linking nearby gyri in SWM may represent an overlooked region of WM changes in Alzheimer’s disease that could help maximise the usefulness of WM as a neuroimaging biomarker.

## Acknowledgements

The authors would like to thank all research participants who made this study possible, as well as Alzheimer’s Research UK and Iceland Foods Charitable Foundation for funding the Young-Onset Alzheimer’s disease study. The Dementia Research Centre is supported by Alzheimer’s Research UK, Brain Research Trust, and The Wolfson Foundation. They also thank Kirsty Lu, Amelia Carton, Timothy Shakespeare, Keir Yong, Aida Suarez Gonzalez and Silvia Primativo for assistance with neuropsychology assessments. The authors would like to thank Fakhereh Movahedian Attar, Evgeniya Kirilina and Nikolaus Weiskopf for enlightening discussions on superficial white matter.

## Abbreviations

AMICO: Accelerated Microstructural Imaging via Convex Optimisation
AME: Average Marginal Effects
DWM: Deep White Matter
dMRI: diffusion MRI
DTI: Diffusion Tensor Imaging
FDR: False Discovery Rate
FWF: Free Water Fraction
GM: Grey Matter
MD: Mean Diffusivity
MMSE: Mini Mental State Examination
NODDI: Neurite Orientation Dispersion and Density Imaging
NDI: Neurite Density Index
ODI: Orientation Dispersion Index
ROI: Region Of Interest
SWM: Superficial White Matter
TF: Tissue Fraction
WM: White matter

