## Supplementary Material for "Loss and Reorganisation of Superficial White Matter in Alzheimer’s disease: A Diffusion MRI study"

### Linear Mixed Effect Models

#### Model

Below is model used to fit the data to each diffusion metric (FA, MD, NDI, ODI and TF) in each ROI (n = 15). Random effects were separated by group, with random intercepts and slopes allowed to be correlated (i.e. unstructured covariance structure) for the majority of models:

$$y_{ijk} = (\beta_{0i} + b_{0ij}) + (\beta_{1i} + b_{1ij})d_k + \beta_{2i}d_k^2 + \beta_3 c_{ij} + \epsilon_{ijk}$$

$$\begin{pmatrix} b_{0ij} \\ b_{1ij} \end{pmatrix} \sim N\left(0, \begin{pmatrix} \sigma_{0i}^2 & \sigma_{01i} \\ \sigma_{01i} & \sigma_{1i}^2 \end{pmatrix}\right), \epsilon_{ijk} \sim N(0, \sigma_i^2)$$

Where, for the $k$th ($k$=1,2,3,4) measurement for the $j$th participant ($j$ = 1,..,$n_i$) in the $i$th group ($i$=1 YOAD, $i$=2 control):

$y_{ijk}$ = dMRI metric
$d_k$ = Fixed distance (mm) sampled from the GM/WM boundary. Moving from GM towards the SWM/DWM:

- GM (+1mm) was set as $d_1$ =0
- GM/WM boundary (0mm) as $d_2$ =1
- SWM (-1mm) as $d_3$ =2
- SWM/DWM (-2mm) as $d_4$ =3

$c_{ij}$ = cortical thickness (mm) as a constant over $k$
$\beta_{0i}$ = Fixed intercepts for the two groups
$\beta_{1i}$ = Fixed linear effects for the two groups
$\beta_{2i}$ = Fixed quadratic effects for the two groups
$\beta_3$ = Fixed cortical thickness effect
$b_{0ij}$ = Random intercept for the $j$th participant in the $i$th group
$b_{1ij}$ = Random slope for the $j$th participant in the $i$th group
$\epsilon_{ijk}$ = Residual

In regions and dMRI metrics where the model was too complex (6/75 models), we fit the same model but removed the control group's random slope.

#### Example R code for fitting models

The above model was implemented in R using the `lme4` library. Separate models were run for each combination of dMRI metric and ROI. Group-specific random effects were achieved by creating separate intercept and distance vectors (e.g. `(0 + control + distance_control | Participant)`). Below shows example code for modelling NDI measures across the GM/WM boundary in the parahippocampal cortex (`phippho_ndi`) where random intercepts and slopes are included for both control and YOAD groups.

```
# Set tolerance for convergence to resolve convergence problems at cost of slower fit (see lme4 `?convergence` he
lp file)
strict_tol <- lmerControl(optCtrl=list(xtol_abs=1e-8, ftol_abs=1e-8))

# Run model
phippo_ndi_model <- lmer(DWI ~ distance + I(distance^2)+ group + distance*group + I(distance^2)*group + cortical_
thick +
                                    (0 + control + distance_control | Participant) +
                                    (0 + AD + distance_AD | Participant),
                                  data=phippo_ndi,
                                  REML=TRUE,
                                  control = strict_tol)
# View model results
summary(phippo_ndi_model)
```

#### Extracting Average Marginal Effects (AMEs)

Marginal effects were estimated from the LME models to assess differences between YOAD and control groups at each sampled distance along the cortical profile. This was done using the `margins` library which emulates the average marginal effects command used in STATA.

```
# Extract marginal effects at each distance (0:3 = 1mm to -2mm) between groups (0 = control, 1 = YOAD)
phippo_ndi_marg <- as.data.frame(summary(margins(phippo_ndi_model, at = list(distance = 0:3, group = 0:1))))

# View marginal effects between groups
phippo_ndi_marg %>% filter(factor == "group")
```

The output provides the average marginal effect (AME) between groups at each distance, standard error, z value and p value. All AME p-values were then adjusted using false discovery rate (FDR) using the `p.adjust` package, where pFDR < 0.05 indicates statistical significance at our chosen threshold of 0.05 after correcting for multiple comparisons.

### Average Marginal Effects

#### Plots

##### Supplementary Figure 1: DTI AMEs for all ROIs Along the Cortical Profile

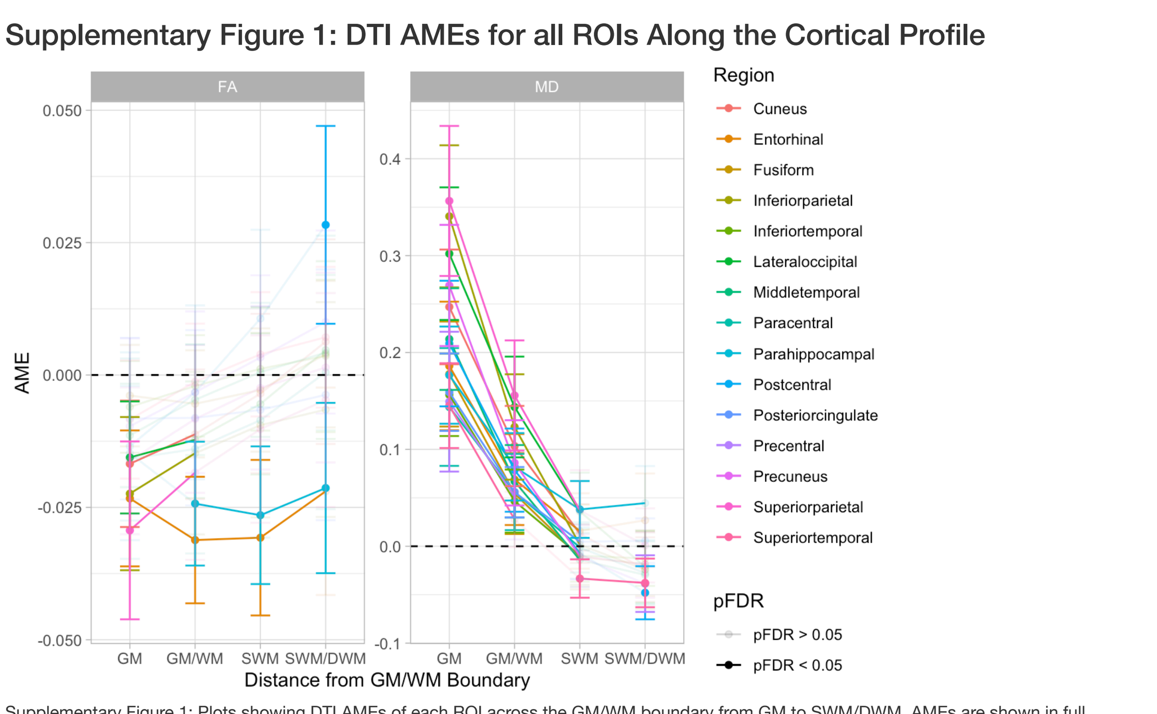


Supplementary Figure 1: Plots showing DTI AMEs of each ROI across the GM/WM boundary from GM to SWM/DWM. AMEs are shown in full colour when pFDR < 0.05 and transparent when pFDR > 0.05.

##### Supplementary Figure 2: NODDI AMEs for all ROIs Along the Cortical Profile

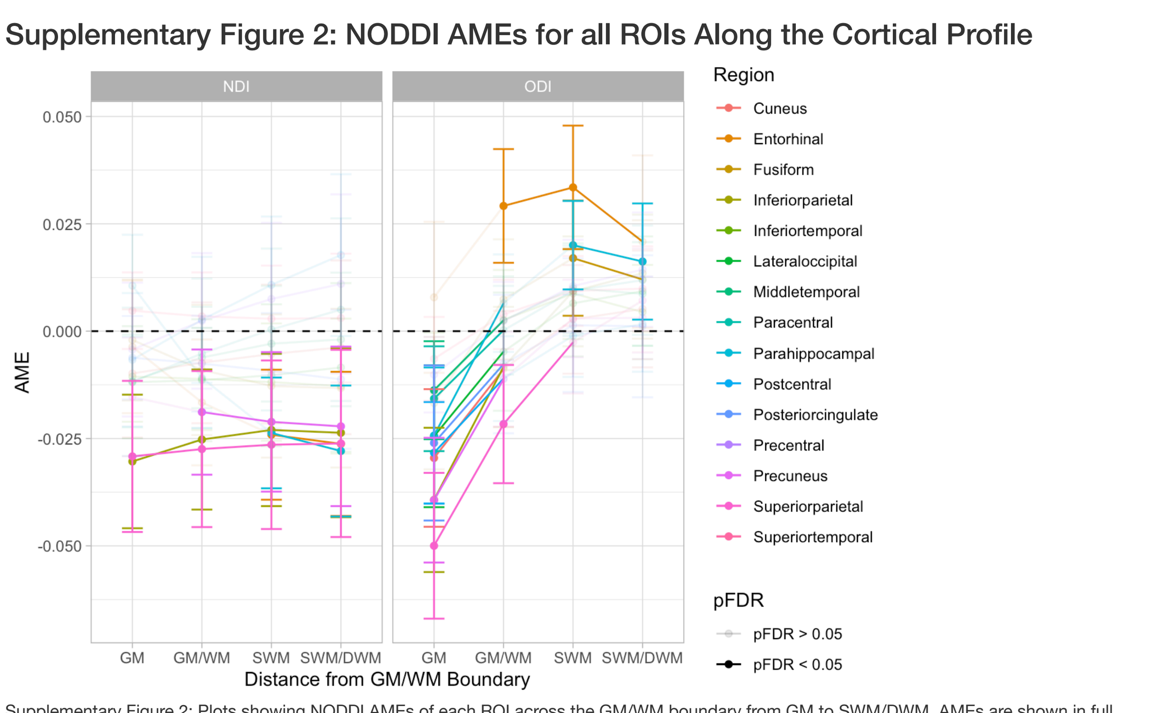


Supplementary Figure 2: Plots showing NODDI AMEs of each ROI across the GM/WM boundary from GM to SWM/DWM. AMEs are shown in full colour when pFDR < 0.05 and transparent when pFDR > 0.05.

##### Supplementary Figure 3: TF AMEs for all ROIs Along the Cortical Profile

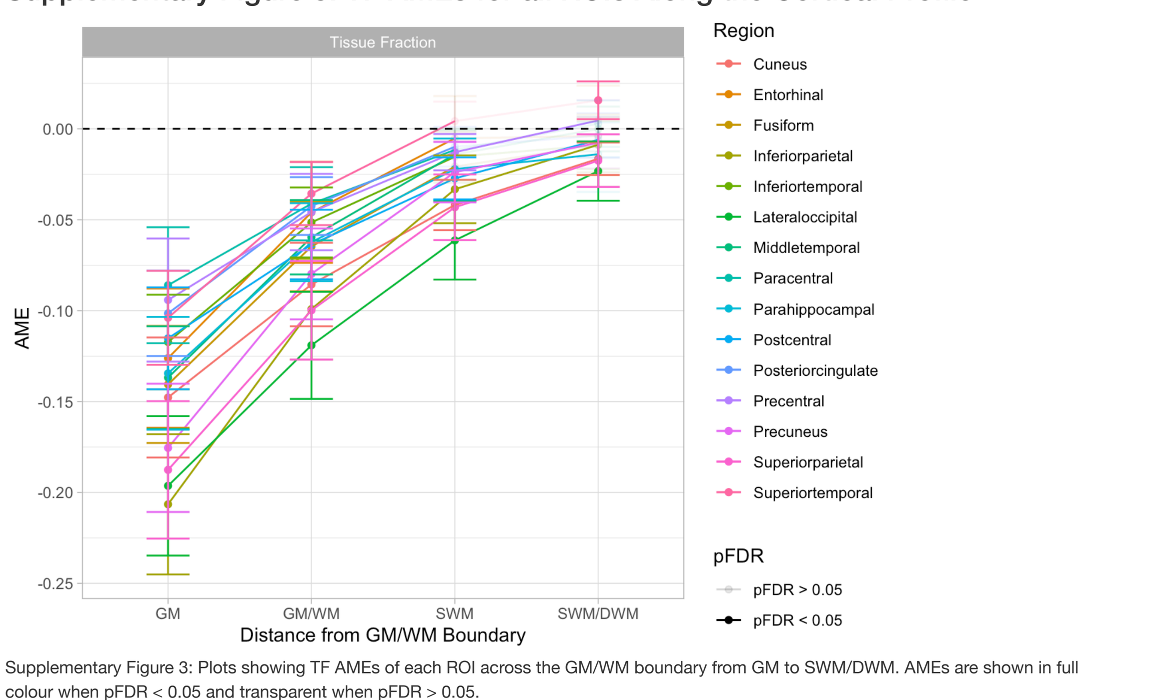


Supplementary Figure 3: Plots showing TF AMEs of each ROI across the GM/WM boundary from GM to SWM/DWM. AMEs are shown in full colour when pFDR < 0.05 and transparent when pFDR > 0.05.

#### Tables

##### Supplementary Table 1: DTI AMEs for all ROIs and Distances

| ROI | Measure | GM | GM/WM | SWM | SWM/DWM |
|---|---|---|---|---|---|
| Cuneus | FA | -0.017 [-0.029, -0.005] | -0.011 [-0.023, 0.001] | -0.004 [-0.019, 0.012] | 0.006 [-0.013, 0.026] |
| | MD | 0.247 [0.188, 0.306] | 0.103 [0.062, 0.145] | 0.013 [-0.017, 0.043] | -0.024 [-0.057, 0.009] |
| Entorhinal | FA | -0.023 [-0.036, -0.010] | -0.031 [-0.043, -0.019] | -0.031 [-0.045, -0.016] | -0.022 [-0.042, -0.002] |
| | MD | 0.186 [0.120, 0.252] | 0.069 [0.022, 0.116] | 0.016 [-0.023, 0.055] | 0.027 [-0.021, 0.075] |
| Fusiform | FA | -0.004 [-0.013, 0.006] | -0.005 [-0.016, 0.005] | -0.003 [-0.014, 0.009] | 0.004 [-0.010, 0.018] |
| | MD | 0.178 [0.124, 0.232] | 0.054 [0.013, 0.096] | -0.011 [-0.045, 0.023] | -0.018 [-0.051, 0.015] |
| Inferiorparietal | FA | -0.022 [-0.037, -0.008] | -0.015 [-0.030, 0.001] | -0.009 [-0.027, 0.008] | -0.007 [-0.027, 0.014] |
| | MD | 0.341 [0.267, 0.414] | 0.123 [0.069, 0.178] | 0.002 [-0.040, 0.043] | -0.024 [-0.065, 0.017] |
| Inferiortemporal | FA | -0.006 [-0.015, 0.003] | -0.002 [-0.012, 0.008] | 0.001 [-0.010, 0.013] | 0.004 [-0.011, 0.018] |
| | MD | 0.156 [0.114, 0.199] | 0.047 [0.014, 0.079] | -0.010 [-0.037, 0.017] | -0.013 [-0.041, 0.016] |
| Lateraloccipital | FA | -0.016 [-0.026, -0.005] | -0.012 [-0.023, -0.001] | -0.005 [-0.019, 0.008] | 0.005 [-0.012, 0.021] |
| | MD | 0.302 [0.234, 0.370] | 0.144 [0.092, 0.196] | 0.036 [-0.004, 0.076] | -0.021 [-0.058, 0.016] |
| Middletemporal | FA | -0.011 [-0.021, -0.002] | -0.005 [-0.015, 0.006] | 0.001 [-0.012, 0.013] | 0.004 [-0.010, 0.019] |
| | MD | 0.214 [0.161, 0.266] | 0.067 [0.030, 0.104] | -0.014 [-0.042, 0.014] | -0.030 [-0.059, 0.000] |
| Paracentral | FA | -0.016 [-0.035, 0.003] | -0.014 [-0.034, 0.006] | -0.009 [-0.031, 0.014] | 0.001 [-0.025, 0.026] |
| | MD | 0.144 [0.083, 0.205] | 0.056 [0.017, 0.095] | -0.001 [-0.027, 0.025] | -0.028 [-0.061, 0.005] |
| Parahippocampal | FA | -0.015 [-0.027, -0.002] | -0.024 [-0.036, -0.013] | -0.026 [-0.039, -0.013] | -0.021 [-0.037, -0.005] |
| | MD | 0.177 [0.126, 0.227] | 0.082 [0.047, 0.117] | 0.038 [0.009, 0.067] | 0.045 [0.006, 0.083] |
| Postcentral | FA | -0.013 [-0.031, 0.004] | -0.003 [-0.020, 0.013] | 0.011 [-0.006, 0.027] | 0.028 [0.010, 0.047] |
| | MD | 0.209 [0.144, 0.274] | 0.079 [0.036, 0.121] | -0.007 [-0.034, 0.019] | -0.048 [-0.075, -0.021] |
| Posteriorcingulate | FA | -0.008 [-0.024, 0.007] | -0.008 [-0.025, 0.008] | -0.007 [-0.026, 0.013] | -0.004 [-0.027, 0.020] |
| | MD | 0.159 [0.119, 0.199] | 0.056 [0.030, 0.082] | 0.005 [-0.013, 0.023] | 0.006 [-0.017, 0.029] |
| Precentral | FA | -0.010 [-0.026, 0.007] | -0.003 [-0.019, 0.012] | 0.003 [-0.012, 0.019] | 0.010 [-0.007, 0.027] |
| | MD | 0.149 [0.077, 0.221] | 0.052 [0.007, 0.096] | -0.011 [-0.034, 0.013] | -0.039 [-0.068, -0.009] |
| Precuneus | FA | -0.016 [-0.030, -0.002] | -0.008 [-0.022, 0.006] | -0.002 [-0.018, 0.013] | 0.001 [-0.017, 0.019] |
| | MD | 0.269 [0.207, 0.332] | 0.086 [0.042, 0.130] | -0.010 [-0.042, 0.022] | -0.019 [-0.053, 0.015] |
| Superiorparietal | FA | -0.029 [-0.046, -0.013] | -0.018 [-0.035, -0.002] | -0.010 [-0.028, 0.007] | -0.005 [-0.025, 0.015] |
| | MD | 0.356 [0.279, 0.434] | 0.156 [0.099, 0.213] | 0.037 [-0.004, 0.079] | 0.001 [-0.037, 0.039] |
| Superiortemporal | FA | -0.008 [-0.020, 0.003] | -0.001 [-0.012, 0.010] | 0.004 [-0.008, 0.016] | 0.007 [-0.006, 0.020] |
| | MD | 0.145 [0.101, 0.189] | 0.028 [-0.001, 0.056] | -0.033 [-0.053, -0.014] | -0.038 [-0.063, -0.013] |

*Note:*
Values reflect Average Marginal Effects (AME) and associated 95% confidence intervals.
Colours indicate FDR adjusted p-value thresholds. Red = pFDR < 0.001, orange = pFDR < 0.01, blue = pFDR < 0.05

##### Supplementary Table 2: NODDI AMEs for all ROIs and Distances

| ROI | Measure | GM | GM/WM | SWM | SWM/DWM |
|---|---|---|---|---|---|
| Cuneus | NDI | -0.010 [-0.025, 0.005] | -0.007 [-0.021, 0.007] | -0.005 [-0.021, 0.011] | -0.004 [-0.024, 0.016] |
| | ODI | -0.030 [-0.046, -0.014] | -0.009 [-0.022, 0.005] | 0.003 [-0.010, 0.015] | 0.005 [-0.008, 0.019] |
| Entorhinal | NDI | -0.004 [-0.019, 0.012] | -0.017 [-0.031, -0.002] | -0.024 [-0.039, -0.009] | -0.026 [-0.043, -0.009] |
| | ODI | 0.008 [-0.010, 0.025] | 0.029 [0.016, 0.042] | 0.033 [0.019, 0.048] | 0.021 [0.001, 0.041] |
| Fusiform | NDI | -0.002 [-0.016, 0.012] | -0.009 [-0.024, 0.006] | -0.013 [-0.029, 0.004] | -0.013 [-0.032, 0.005] |
| | ODI | -0.017 [-0.032, -0.001] | 0.007 [-0.007, 0.021] | 0.017 [0.004, 0.030] | 0.012 [-0.002, 0.026] |
| Inferiorparietal | NDI | -0.030 [-0.046, -0.015] | -0.025 [-0.042, -0.009] | -0.023 [-0.041, -0.005] | -0.024 [-0.043, -0.004] |
| | ODI | -0.039 [-0.056, -0.022] | -0.008 [-0.022, 0.006] | 0.009 [-0.004, 0.022] | 0.014 [0.000, 0.027] |
| Inferiortemporal | NDI | -0.011 [-0.021, 0.000] | -0.011 [-0.023, 0.001] | -0.012 [-0.026, 0.002] | -0.013 [-0.029, 0.003] |
| | ODI | -0.014 [-0.028, 0.000] | 0.003 [-0.009, 0.014] | 0.009 [-0.002, 0.019] | 0.004 [-0.007, 0.015] |
| Lateraloccipital | NDI | -0.012 [-0.025, 0.001] | -0.011 [-0.026, 0.003] | -0.010 [-0.027, 0.006] | -0.008 [-0.028, 0.011] |
| | ODI | -0.024 [-0.041, -0.008] | -0.005 [-0.019, 0.009] | 0.006 [-0.006, 0.019] | 0.009 [-0.003, 0.022] |
| Middletemporal | NDI | -0.012 [-0.022, -0.001] | -0.006 [-0.018, 0.006] | -0.003 [-0.016, 0.011] | -0.002 [-0.017, 0.014] |
| | ODI | -0.014 [-0.025, -0.002] | 0.003 [-0.008, 0.013] | 0.010 [0.000, 0.020] | 0.009 [-0.003, 0.021] |
| Paracentral | NDI | -0.012 [-0.029, 0.005] | -0.005 [-0.023, 0.012] | 0.000 [-0.018, 0.019] | 0.005 [-0.016, 0.026] |
| | ODI | -0.016 [-0.028, -0.004] | 0.000 [-0.010, 0.011] | 0.009 [-0.001, 0.020] | 0.012 [-0.001, 0.025] |
| Parahippocampal | NDI | 0.011 [-0.001, 0.022] | -0.011 [-0.023, 0.001] | -0.024 [-0.037, -0.011] | -0.028 [-0.043, -0.013] |
| | ODI | -0.024 [-0.040, -0.008] | 0.007 [-0.005, 0.018] | 0.020 [0.010, 0.030] | 0.016 [0.003, 0.030] |
| Postcentral | NDI | -0.007 [-0.022, 0.009] | 0.003 [-0.012, 0.017] | 0.011 [-0.005, 0.027] | 0.018 [-0.001, 0.037] |
| | ODI | -0.028 [-0.040, -0.017] | -0.011 [-0.021, -0.001] | -0.001 [-0.011, 0.009] | 0.002 [-0.009, 0.013] |
| Posteriorcingulate | NDI | -0.006 [-0.016, 0.004] | -0.008 [-0.019, 0.004] | -0.009 [-0.023, 0.004] | -0.011 [-0.027, 0.005] |
| | ODI | -0.026 [-0.044, -0.008] | -0.008 [-0.024, 0.008] | 0.001 [-0.014, 0.017] | 0.001 [-0.015, 0.018] |
| Precentral | NDI | -0.004 [-0.020, 0.011] | 0.002 [-0.013, 0.018] | 0.007 [-0.010, 0.025] | 0.011 [-0.010, 0.032] |
| | ODI | -0.010 [-0.019, -0.001] | 0.002 [-0.007, 0.011] | 0.011 [0.000, 0.021] | 0.014 [0.001, 0.028] |
| Precuneus | NDI | -0.015 [-0.029, -0.002] | -0.019 [-0.033, -0.004] | -0.021 [-0.037, -0.005] | -0.022 [-0.041, -0.004] |
| | ODI | -0.039 [-0.054, -0.025] | -0.011 [-0.022, 0.000] | 0.003 [-0.006, 0.012] | 0.003 [-0.006, 0.013] |
| Superiorparietal | NDI | -0.029 [-0.047, -0.012] | -0.027 [-0.046, -0.009] | -0.026 [-0.046, -0.007] | -0.026 [-0.048, -0.004] |
| | ODI | -0.050 [-0.067, -0.033] | -0.022 [-0.035, -0.008] | -0.003 [-0.015, 0.009] | 0.007 [-0.005, 0.019] |
| Superiortemporal | NDI | 0.005 [-0.004, 0.014] | 0.003 [-0.007, 0.014] | 0.003 [-0.009, 0.015] | 0.003 [-0.012, 0.018] |
| | ODI | -0.006 [-0.016, 0.003] | 0.004 [-0.004, 0.012] | 0.009 [0.001, 0.017] | 0.010 [0.000, 0.020] |

*Note:*
Values reflect Average Marginal Effects (AME) and associated 95% confidence intervals.
Colours indicate FDR adjusted p-value thresholds. Red = pFDR < 0.001, orange = pFDR < 0.01, blue = pFDR < 0.05

##### Supplementary Table 3: Tissue Fraction AMEs for all ROIs and Distances

| ROI | Measure | GM | GM/WM | SWM | SWM/DWM |
|---|---|---|---|---|---|
| Cuneus | Tissue Fraction | -0.148 [-0.181, -0.115] | -0.086 [-0.109, -0.063] | -0.042 [-0.056, -0.028] | -0.016 [-0.025, -0.008] |
| Entorhinal | | -0.126 [-0.164, -0.088] | -0.046 [-0.074, -0.018] | -0.005 [-0.028, 0.018] | -0.004 [-0.032, 0.024] |
| Fusiform | | -0.141 [-0.173, -0.108] | -0.065 [-0.089, -0.040] | -0.021 [-0.040, -0.002] | -0.009 [-0.026, 0.008] |
| Inferiorparietal | | -0.207 [-0.245, -0.168] | -0.099 [-0.127, -0.071] | -0.033 [-0.052, -0.015] | -0.009 [-0.024, 0.006] |
| Inferiortemporal | | -0.117 [-0.143, -0.091] | -0.052 [-0.071, -0.032] | -0.015 [-0.030, -0.001] | -0.009 [-0.022, 0.004] |
| Lateraloccipital | | -0.196 [-0.235, -0.158] | -0.119 [-0.149, -0.090] | -0.061 [-0.083, -0.040] | -0.023 [-0.040, -0.007] |
| Middletemporal | | -0.137 [-0.165, -0.109] | -0.060 [-0.080, -0.039] | -0.014 [-0.028, 0.000] | 0.000 [-0.012, 0.012] |
| Paracentral | | -0.086 [-0.118, -0.054] | -0.041 [-0.061, -0.021] | -0.012 [-0.023, -0.001] | 0.003 [-0.010, 0.016] |
| Parahippocampal | | -0.135 [-0.166, -0.103] | -0.062 [-0.084, -0.041] | -0.022 [-0.039, -0.005] | -0.014 [-0.035, 0.007] |
| Postcentral | | -0.115 [-0.143, -0.087] | -0.064 [-0.083, -0.045] | -0.027 [-0.039, -0.016] | -0.006 [-0.016, 0.004] |
| Posteriorcingulate | | -0.102 [-0.125, -0.078] | -0.042 [-0.058, -0.027] | -0.010 [-0.021, 0.001] | -0.004 [-0.016, 0.009] |
| Precentral | | -0.094 [-0.128, -0.060] | -0.046 [-0.067, -0.025] | -0.013 [-0.023, -0.003] | 0.005 [-0.007, 0.016] |
| Precuneus | | -0.175 [-0.211, -0.140] | -0.080 [-0.105, -0.055] | -0.024 [-0.040, -0.007] | -0.008 [-0.022, 0.006] |
| Superiorparietal | | -0.188 [-0.225, -0.150] | -0.100 [-0.127, -0.073] | -0.043 [-0.061, -0.025] | -0.017 [-0.032, -0.003] |
| Superiortemporal | | -0.104 [-0.130, -0.078] | -0.036 [-0.053, -0.018] | 0.004 [-0.006, 0.015] | 0.016 [0.005, 0.026] |

*Note:*
Values reflect Average Marginal Effects (AME) and associated 95% confidence intervals.
Colours indicate FDR adjusted p-value thresholds. Red = pFDR < 0.001, orange = pFDR < 0.01, blue = pFDR < 0.05

##### Supplementary Figure 4: AMEs with Uncorrected p-values < 0.05

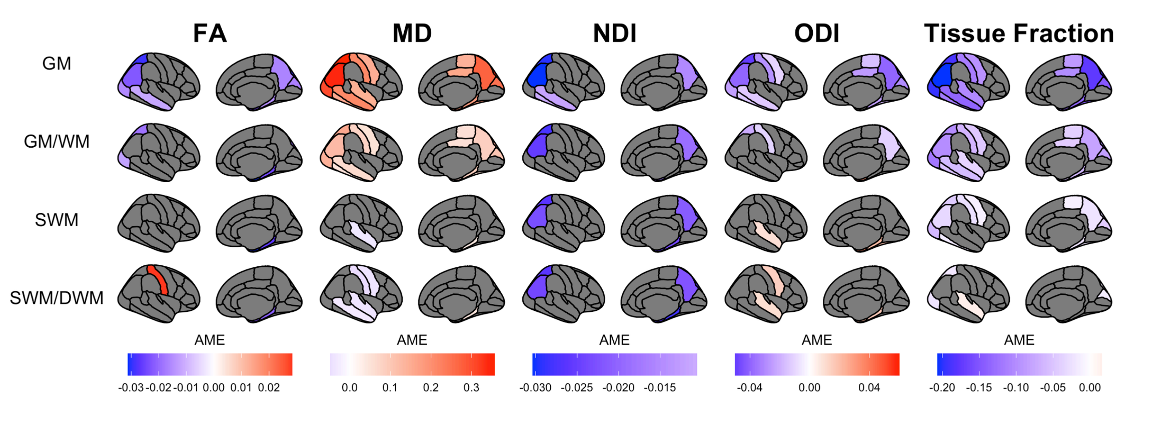


Supplementary Figure 4: Plots showing AMEs along the profile for each dMRI metric. Visible AMEs are significant at p < 0.05 and not corrected for multiple comparisons.

### Conventional Mean ROI NODDI values

The supplementary results below show AMEs that were obtained when we did not perform tissue-weighted averages of ROIs described in the main text (see *Regions of Interest (ROIs)* section). NDI and ODI metrics were first masked by ISO to retain voxels with mostly tissue (i.e. only including voxels with < 0.3 ISO), then the same cortical profile extraction and linear mixed effect model analyses were performed. A similar pattern along the cortical profile as observed when using tissue-weighted averages in the main text can be observed in the SWM: overall lower NDI in YOAD individuals compared to controls, and overall higher ODI in YOAD individuals compared to controls. However, group differences are greater in magnitude and extent within the cortical GM when compared to using tissue-weighted average NODDI measures.

##### Supplementary Figure 5: Conventional Mean ROI NODDI AMEs for all ROIs Along the Cortical Profile

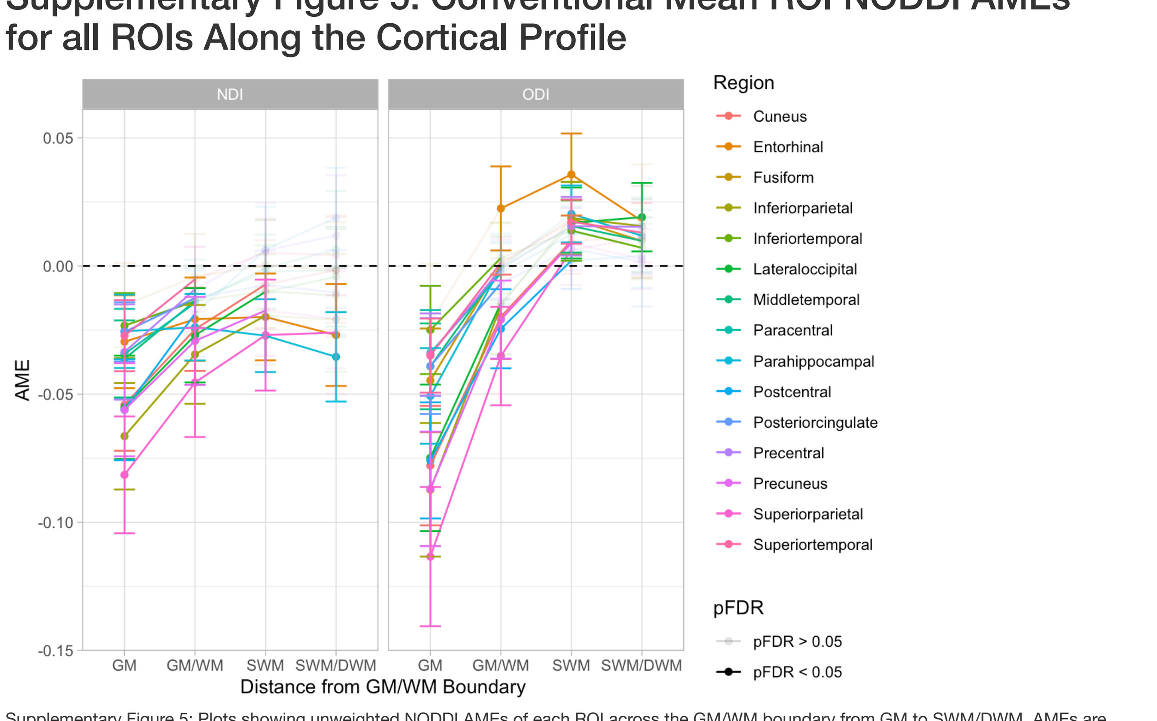


Supplementary Figure 5: Plots showing unweighted NODDI AMEs of each ROI across the GM/WM boundary from GM to SWM/DWM. AMEs are shown in full colour when pFDR < 0.05 and transparent when pFDR > 0.05.

##### Supplementary Table 4: Conventional Mean ROI NODDI AMEs for all ROIs and Distances

| ROI | Measure | GM | GM/WM | SWM | SWM/DWM |
|---|---|---|---|---|---|
| Cuneus | NDI | -0.054 [-0.072, -0.036] | -0.025 [-0.041, -0.009] | -0.007 [-0.024, 0.010] | -0.002 [-0.022, 0.019] |
| | ODI | -0.078 [-0.101, -0.055] | -0.020 [-0.036, -0.003] | 0.010 [-0.003, 0.023] | 0.011 [-0.004, 0.026] |
| Entorhinal | NDI | -0.030 [-0.048, -0.011] | -0.021 [-0.037, -0.005] | -0.020 [-0.037, -0.003] | -0.027 [-0.047, -0.007] |
| | ODI | -0.022 [-0.045, 0.001] | 0.022 [0.006, 0.039] | 0.036 [0.020, 0.052] | 0.018 [-0.005, 0.040] |
| Fusiform | NDI | -0.015 [-0.032, 0.001] | -0.004 [-0.021, 0.012] | 0.000 [-0.017, 0.018] | -0.002 [-0.021, 0.017] |
| | ODI | -0.045 [-0.065, -0.024] | 0.000 [-0.016, 0.017] | 0.018 [0.004, 0.033] | 0.010 [-0.005, 0.025] |
| Inferiorparietal | NDI | -0.066 [-0.087, -0.046] | -0.035 [-0.054, -0.015] | -0.019 [-0.038, 0.000] | -0.021 [-0.041, -0.001] |
| | ODI | -0.087 [-0.113, -0.061] | -0.016 [-0.034, 0.003] | 0.019 [0.005, 0.033] | 0.016 [0.000, 0.031] |
| Inferiortemporal | NDI | -0.023 [-0.036, -0.011] | -0.014 [-0.027, -0.001] | -0.010 [-0.024, 0.005] | -0.012 [-0.028, 0.005] |
| | ODI | -0.025 [-0.042, -0.008] | 0.003 [-0.011, 0.017] | 0.014 [0.002, 0.026] | 0.007 [-0.005, 0.019] |
| Lateraloccipital | NDI | -0.055 [-0.075, -0.035] | -0.027 [-0.045, -0.009] | -0.010 [-0.028, 0.008] | -0.004 [-0.024, 0.015] |
| | ODI | -0.075 [-0.103, -0.046] | -0.015 [-0.035, 0.006] | 0.017 [0.003, 0.031] | 0.019 [0.006, 0.032] |
| Middletemporal | NDI | -0.036 [-0.051, -0.021] | -0.014 [-0.028, 0.000] | -0.002 [-0.017, 0.012] | -0.002 [-0.018, 0.014] |
| | ODI | -0.039 [-0.056, -0.022] | -0.001 [-0.013, 0.012] | 0.016 [0.005, 0.026] | 0.010 [-0.002, 0.022] |
| Paracentral | NDI | -0.034 [-0.052, -0.017] | -0.014 [-0.031, 0.002] | -0.001 [-0.020, 0.018] | 0.006 [-0.016, 0.029] |
| | ODI | -0.034 [-0.051, -0.017] | -0.003 [-0.015, 0.009] | 0.012 [0.001, 0.023] | 0.012 [-0.003, 0.027] |
| Parahippocampal | NDI | -0.025 [-0.040, -0.011] | -0.024 [-0.037, -0.011] | -0.027 [-0.041, -0.013] | -0.035 [-0.053, -0.018] |
| | ODI | -0.051 [-0.069, -0.032] | -0.001 [-0.014, 0.013] | 0.020 [0.009, 0.031] | 0.012 [-0.002, 0.026] |
| Postcentral | NDI | -0.056 [-0.076, -0.036] | -0.019 [-0.035, -0.002] | 0.006 [-0.010, 0.023] | 0.019 [-0.001, 0.038] |
| | ODI | -0.076 [-0.099, -0.053] | -0.025 [-0.040, -0.009] | 0.002 [-0.009, 0.013] | 0.004 [-0.008, 0.016] |
| Posteriorcingulate | NDI | -0.026 [-0.037, -0.014] | -0.013 [-0.024, -0.001] | -0.007 [-0.021, 0.006] | -0.010 [-0.027, 0.006] |
| | ODI | -0.039 [-0.058, -0.020] | -0.007 [-0.023, 0.010] | 0.007 [-0.009, 0.023] | 0.002 [-0.016, 0.019] |
| Precentral | NDI | -0.034 [-0.052, -0.015] | -0.009 [-0.026, 0.008] | 0.006 [-0.013, 0.025] | 0.012 [-0.012, 0.035] |
| | ODI | -0.035 [-0.051, -0.019] | -0.001 [-0.013, 0.010] | 0.016 [0.004, 0.027] | 0.015 [-0.001, 0.031] |
| Precuneus | NDI | -0.056 [-0.074, -0.038] | -0.029 [-0.046, -0.012] | -0.017 [-0.035, 0.000] | -0.021 [-0.040, -0.001] |
| | ODI | -0.087 [-0.109, -0.065] | -0.021 [-0.036, -0.006] | 0.009 [-0.002, 0.020] | 0.003 [-0.009, 0.014] |
| Superiorparietal | NDI | -0.081 [-0.104, -0.059] | -0.045 [-0.067, -0.024] | -0.027 [-0.049, -0.005] | -0.026 [-0.050, -0.002] |
| | ODI | -0.113 [-0.141, -0.086] | -0.035 [-0.054, -0.016] | 0.006 [-0.007, 0.020] | 0.011 [-0.003, 0.025] |
| Superiortemporal | NDI | -0.027 [-0.041, -0.013] | -0.005 [-0.018, 0.007] | 0.005 [-0.008, 0.019] | 0.004 [-0.011, 0.020] |
| | ODI | -0.035 [-0.049, -0.020] | 0.001 [-0.009, 0.011] | 0.017 [0.009, 0.026] | 0.013 [0.001, 0.025] |

*Note:*
Values reflect Average Marginal Effects (AME) and associated 95% confidence intervals.
Colours indicate FDR adjusted p-value thresholds. Red = pFDR < 0.001, orange = pFDR < 0.01, blue = pFDR < 0.05

##### Supplementary Figure 6: Conventional Mean ROI NODDI AMEs for NDI and ODI

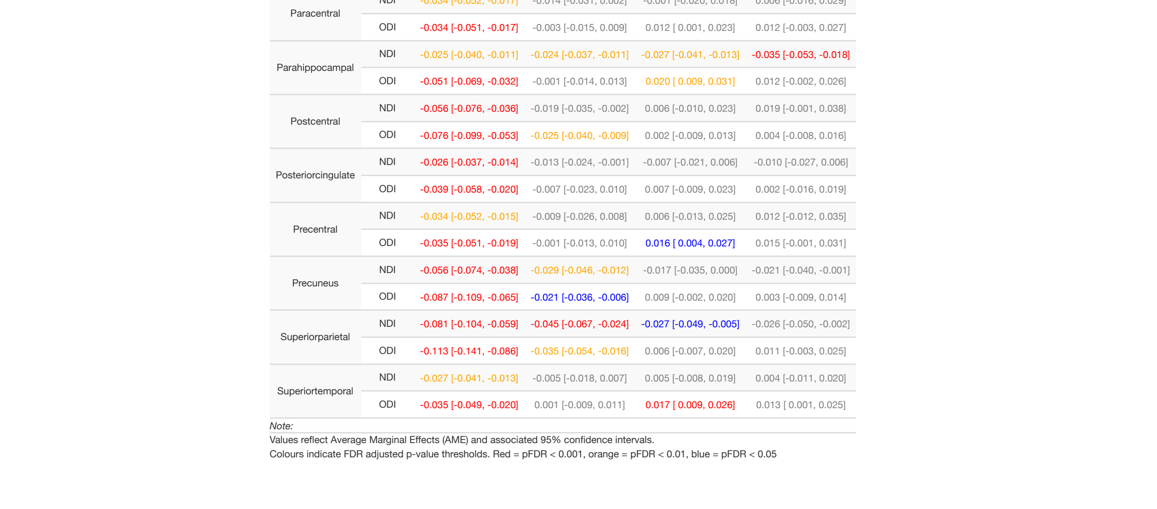


Supplementary Figure 6: Plots showing unweighted NDI and ODI AMEs along the cortical profile. AMEs displayed here are corrected for multiple comparisons (pFDR < 0.05).

### Relationship Between TF and MD

##### Supplementary Figure 7: TF vs MD Along the Cortical Profile in all ROIs

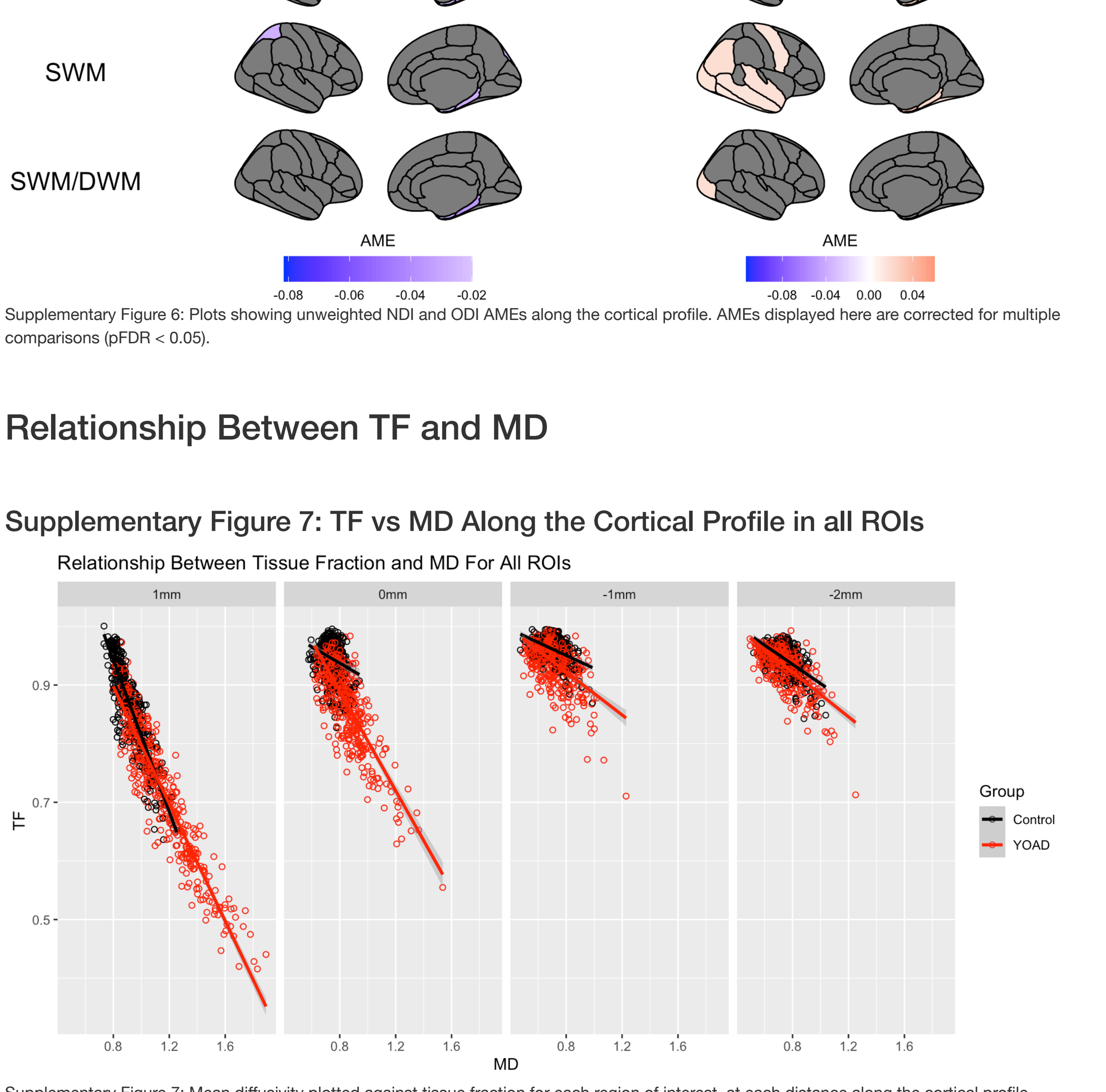


Supplementary Figure 7: Mean diffusivity plotted against tissue fraction for each region of interest, at each distance along the cortical profile (GM=1mm, GM/WM=0mm, SWM=-1mm, SWM/DWM=-2mm). Colours indicate controls (black) and YOAD (red).
